# Machine learning for comprehensive interaction modelling improves disease risk prediction in the UK Biobank

**DOI:** 10.1101/2024.08.07.24311604

**Authors:** Heli Julkunen, Juho Rousu

## Abstract

Understanding how risk factors interact to jointly influence disease risk can provide insights into disease development and improve risk prediction. We introduce *survivalFM*, a machine learning extension to the widely used Cox proportional hazards model that incorporates estimation of all potential pairwise interaction effects on time-to-event outcomes. The method relies on learning a low-rank factorized approximation of the interaction effects, hence overcoming the computational and statistical limitations of fitting these terms in models involving many predictor variables. The resulting model is fully interpretable, providing access to the estimates of both individual effects and the approximated interactions. Comprehensive evaluation of *survivalFM* using the UK Biobank dataset across ten disease examples and a variety of clinical risk factors and omics data modalities shows improved discrimination and reclassification performance (65% and 97.5% of the scenarios tested, respectively). Considering a clinical scenario of cardiovascular risk prediction using predictors from the established QRISK3 model, we further show that the comprehensive interaction modelling adds predictive value beyond the individual and age interaction effects currently included. These results demonstrate that comprehensive modelling of interactions can facilitate advanced insights into disease development and improve risk predictions.

## Introduction

Risk prediction models are needed in modern preventive medicine to identify individuals at high risk of disease before clinical symptoms manifest. The ability to predict disease risk is particularly important in managing complex diseases, such as cardiovascular disease, chronic kidney disease, and diabetes, where early intervention can substantially alter patient outcomes. However, accurately predicting disease risk is challenging due to the inherent complexity of most human diseases, which arise from the interplay of genetic, environmental, and lifestyle factors. Traditional methods in survival analysis, such as the widely used Cox proportional hazards regression [1], assume linear effects of predictor variables on time-to-event outcomes. This assumption may lead to oversimplified models that overlook the complex interplay among predictors, potentially missing important biological insights and limiting risk prediction accuracy.

The accuracy of time-to-event prediction models can be improved by incorporating interaction terms, a well-established concept in epidemiology to assess the joint effects of predictors on outcomes [2, 3]. For instance, interaction terms have been shown to be relevant in cardiovascular disease (CVD) risk prediction, where the effects of other risk factors can vary depending on age [4, 5, 6, 7]. However, incorporating these terms in multivariable prediction models typically requires prior hypotheses about which interactions to include. As the number of potential interaction terms increases quadratically with the number of predictor variables in consideration, inclusion of all potential interactions quickly becomes impractical without targeted hypotheses to guide the selection. Therefore, prior multivariable prediction models have typically been constrained to a restricted set of interaction terms known to alter outcome associations, such as those involving age. This limits the discovery of new, potentially relevant interactions. Another commonly employed strategy is to perform statistical testing of individual interaction terms, but this can miss interactions that only become relevant for prediction in the presence of other variables. This challenge becomes particularly pronounced with modern biomedical datasets, which can contain hundreds of potential predictors. While machine learning survival analysis extensions like random survival forests [8] and deep survival models [9, 10] can capture complex non-linearities and interactions in the underlying data, they often compromise interpretability, which is crucial when the goal is to inform clinical decision-making or to obtain insights into the risk factors underlying disease development.

To enhance possibilities to understand and model the joint effects of risk factors on time-to-event disease outcomes, we here present *survivalFM*, a methodological extension to the Cox proportional hazards model that incorporates estimation of all potential pairwise interaction effects among predictor variables. The method is based on an efficient strategy of learning the interaction effects using a low-rank factorized approximation, a concept taken from factorization machines (FMs) [11] and here applied to survival analysis. *survivalFM* combines the factorization of the interaction effects with an efficient quasi-Newton optimization algorithm, thereby overcoming the computational and statistical challenges of fitting comprehensive interaction effects in time-to-event prediction models involving many variables. The resulting model is fully interpretable, providing access to the estimates of both individual effects and the approximated interactions. We demonstrate the performance of *survivalFM* across various data modalities and disease outcomes using data from the UK Biobank. We further highlight an application in a clinical cardiovascular risk prediction scenario and show that *survivalFM* can learn predictive interaction effects which improve identification of high-risk individuals. While we highlight applications in disease risk prediction, the method is generally applicable to modelling any type of time-to-event outcomes.

## Results

### Overview of *survivalFM*

Figure 1 presents an overview of *survivalFM*. We developed *survivalFM* to estimate all potential pairwise interaction effects among input variables for right-censored survival data, such as time to disease onset. It is based on the widely used proportional hazards model [1] which relates the time until an event occurs to a set of predictor variables through a hazard function of the form:

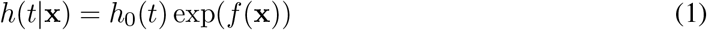

where *h*(*t*|**x**) represents the hazard for an individual at time point *t*, with the baseline hazard function *h*_0_(*t*) describing the time-varying hazard and the partial hazard exp(*f* (**x**)) quantifying the impact of the predictor variables **x** on the baseline hazard. In the standard formulation of the Cox proportional hazards model, the partial hazard exp(*f* (**x**)) is assumed to be parametrized by a linear combination of the predictor variables *f* (**x**) = ***β***^⊤^**x**, with ***β*** giving the weights for the individual variables.

**Figure 1:**
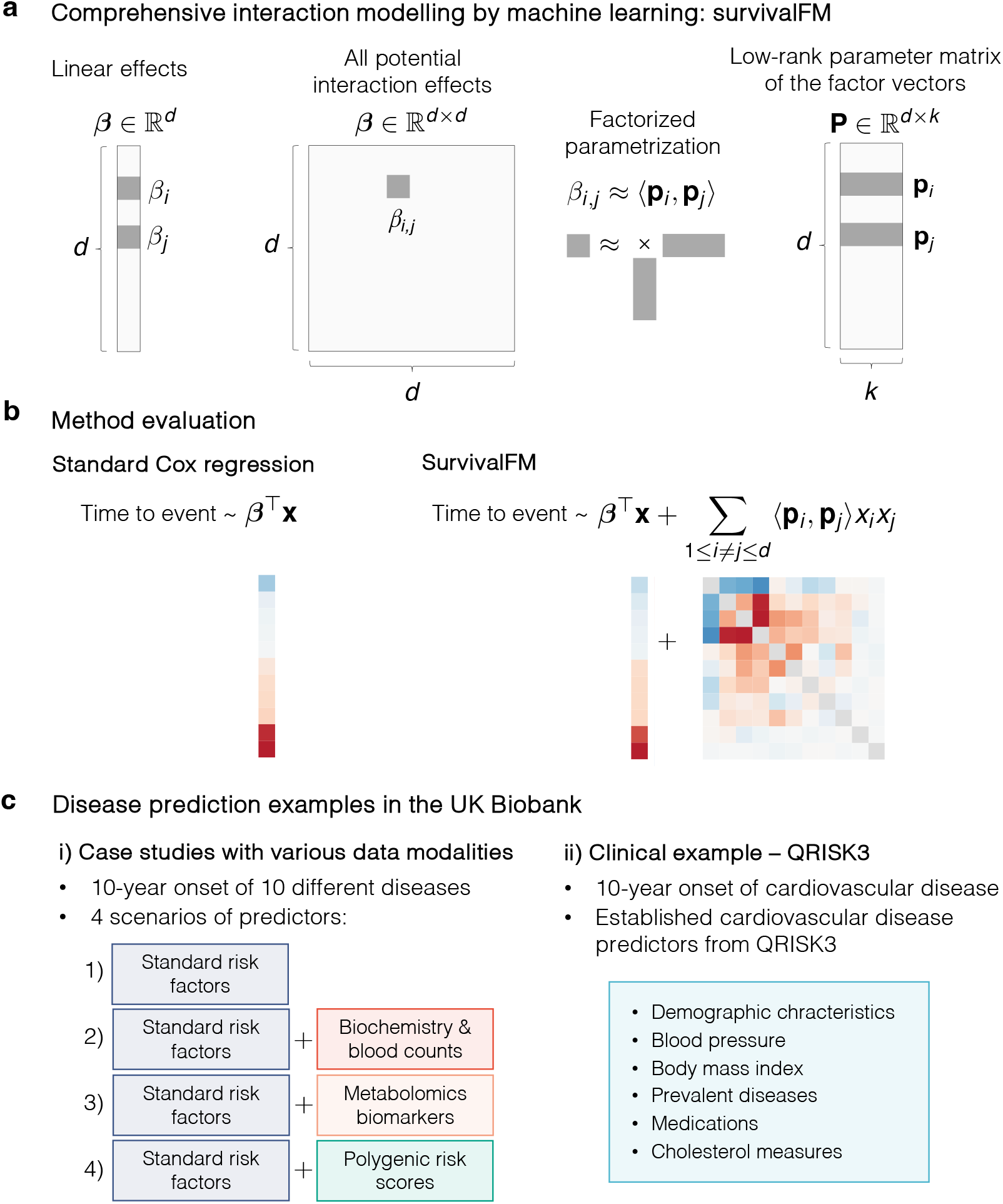
Method overview and evaluation examples. a) A machine learning survival analysis method, *survivalFM*, is developed to estimate linear and all pairwise interaction effects between predictor variables using factorized parametrization of the interaction terms *β*_*i,j*_ ≈ ⟨**p**_*i*_, **p**_*j*_⟩. *d* denotes the number of predictor variables and *k* is a hyperparameter defining the rank of the factorization of the interaction terms. The rank of the factorization is typically much lower than the number of predictor variables (*k* ≪ *d*), enabling computation of the interaction terms even in the presence of many input variables. b) The added value of incorporating comprehensive interaction terms using *survivalFM* is assessed by comparing the performance to the standard linear Cox proportional hazards regression. b) The performance is evaluated in various disease prediction examples: i) case studies with four different predictor sets, each applied to ten disease examples; ii) a clinical example using predictors from the QRISK3 cardiovascular disease (CVD) risk evaluation tool.

In many applications, understanding how variables may interact to jointly impact the hazard rate can provide additional value beyond their independent linear effects. However, directly fitting all potential pairwise interaction terms in a multivariable prediction model quickly becomes challenging due to the quadratic increase in the number of interaction terms as a function of the number of input variables. Hence, we propose *survivalFM*, an extension which adds an approximation of all pairwise interaction effects using a factorized parametrization approach (Figure 1a-b):

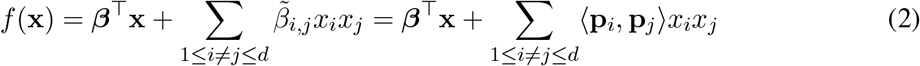

where ⟨·,·⟩ denotes the inner product and *d* denotes the number of predictor variables. The first part contains the linear effects of all predictor variables in the same way as in the standard formulation of the Cox proportional hazards model. The second part contains all pairwise interaction effects between the predictor variables *x*_*i*_ and *x*_*j*_. However, instead of directly estimating the interaction effects *β*_*i,j*_, the factorized parametrization approximates the effects using an inner product between two low-rank latent vectors 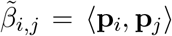. The parameter vectors **p**_*i*_ ∈ ℝ^*k*^ and **p**_*j*_ ∈ ℝ^*k*^ are the row vectors of a low-rank parameter matrix **P** ∈ ℝ^*d*×*k*^ (Figure 1a). Hence, this results in much fewer parameters to estimate, as the rank of the factorization is typically much lower than the total number of predictor variables (*k ≪ d*). With this approach, we avoid the statistical and computational problems that would be encountered with direct estimation of all interactions terms in the presence of many predictor variables, while still maintaining interpretability. The idea of using factorized parametrization strategy originates from factorization machines (FMs) [11], originally proposed for regression and classification tasks in the context of recommender systems. For more details of the model and the fitting procedure, see Methods.

### Study population, disease outcomes and data modalities

To evaluate whether *survivalFM* could improve risk prediction models and provide new insights on the joint effects of risk factors on disease onset, we performed analyses using data from the UK Biobank. This cohort comprises a total of approximately 500,000 participants from the UK, enrolled in 21 recruitment centers across the country. The UK Biobank is renowned for its comprehensive phenotyping and molecular profiling, including routine blood biomarkers and advanced ’omics measurements such as genomics and metabolomics. Baseline characteristics of the study population and a summary of the datasets studied here are summarized in Supplementary Table 1. As disease outcomes, we considered the 10-year incidence of ten example diseases, selected to comprise common diseases and diseases which can benefit from intervention if identified early (Supplementary Tables 2 and 3), excluding participants with a prior record of the disease at baseline.

To assess the performance across different data modalities, we considered four different prediction scenarios that incorporate an array of predictors ranging from traditional clinical predictors to more advanced omics-based data sources (Figure 1c, Methods). In the first scenario, we started from a set of standard cardiovascular risk factors included in the ASCVD risk estimator plus [12], widely recognized in various primary prevention scores. Since these factors have been shown to be predictive beyond cardiovascular diseases [13, 14, 15], we included them as standard risk factors across all analyzed disease examples. We then added sets of more complex data layers to these standard risk factors (Figure 1c). In the second scenario, we added a comprehensive set of hematologic and clinical biochemistry measures to the standard risk factors; in the third scenario, we incorporated a wide range of metabolomic biomarkers, recently shown promise as an assay to inform on multidisease risk [13, 16]; and finally, we included a set of polygenic risk scores for both disease and quantitative traits [17], which have gained interest for their potential to enhance risk prediction models by providing complementary information to traditional risk factors [18, 19, 20].

### *survivalFM* improves risk prediction across various diseases and data modalities

The practical utility of any risk prediction model is determined by its ability to stratify risk and identify high-risk individuals. We evaluated the ability of *survivalFM* to predict future disease risk and benefit from the comprehensive interaction terms by comparing its performance to standard linear Cox proportional hazards regression (Figure 1b), employing L2 (Ridge) regularization in both methods to control model complexity and prevent overfitting (Methods). The performance of the models was evaluated in 10-fold cross-validation, using 20% validation set within each cross-validation cycle to optimize regularization parameters. Analyses were consistently applied across the same sets of predictor variables and fixed cross-validation folds.

By modelling the comprehensive interactions present in the underlying data, *survivalFM* improved the discriminatory performance across a majority of the studied examples as measured by concordance index (C-index; Figure 2). Specifically, statistically significant improvements were noted in 26 of the 40 evaluated scenarios (65%), with a mean improvement in concordance index (ΔC-index) of 0.005. Minor improvements were noted in another 12 out of 40 (30%) of scenarios (mean ΔC-index 0.001). Importantly, none of the studied examples demonstrated a statistically significant decrease in performance with *survivalFM*, highlighting the robustness of *survivalFM*. Absolute values for the C-indices are detailed in Supplementary Table 4, demonstrating good discriminative performance across all models with C-indices in the range 0.72–0.93. Moreover, all models were well calibrated across the UK Biobank cohort (Supplementary Figures 2-5).

**Figure 2:**
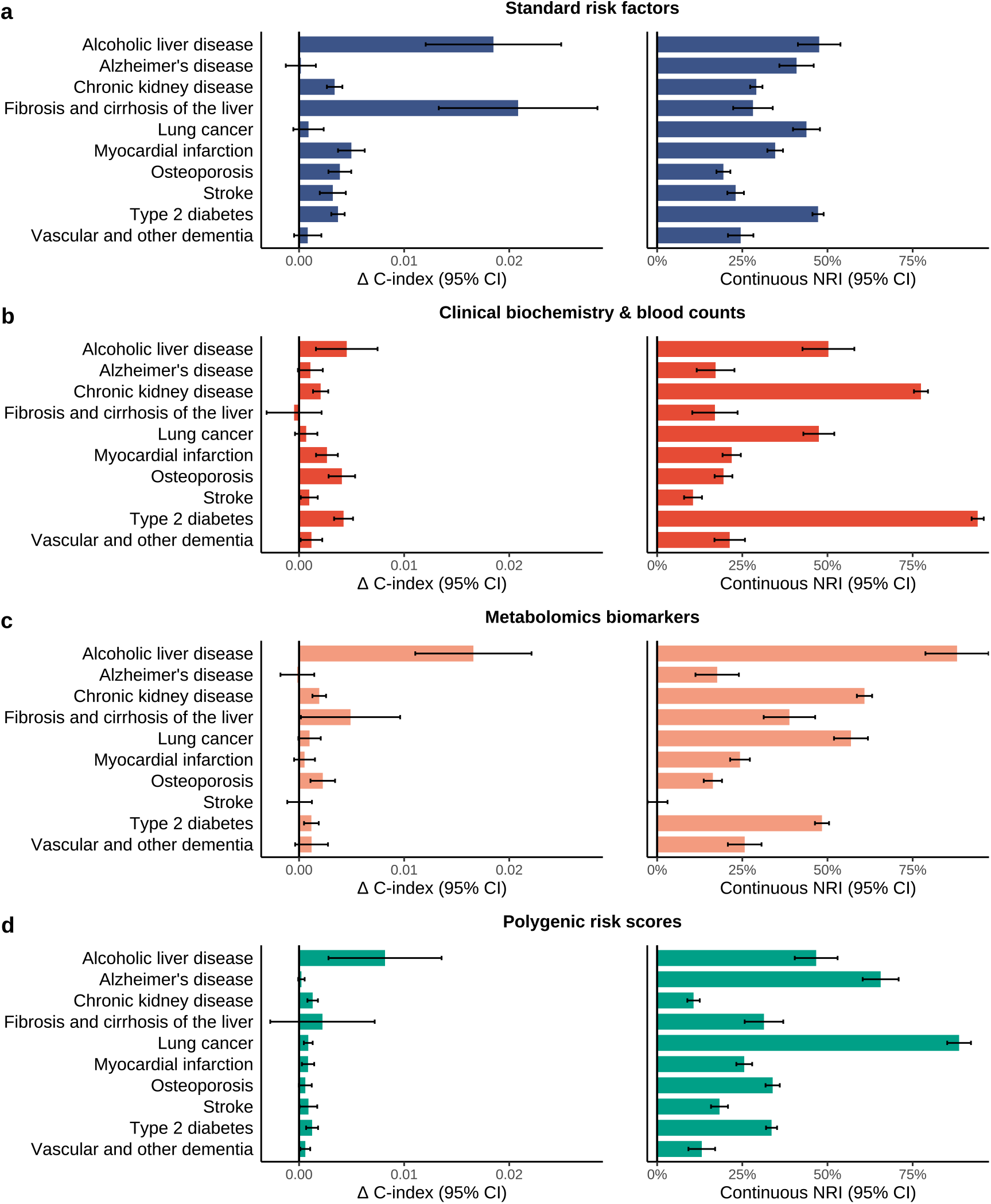
Comprehensive interaction modelling by *survivalFM* improves risk prediction performance across various diseases and data modalities. Comparison of the predictive performance of *survivalFM* to standard linear Cox proportional hazards regression in terms difference in concordance index (Δ C-index) and continuous net reclassification improvement (NRI). Results are shown for ten disease examples (y-axis) across four data modalities: a) standard risk factors (blue; included in all models), b) clinical biochemistry and blood counts (red), c) metabolomics biomarkers (orange) and d) polygenic risk scores (green). Horizontal error bars denote 95% confidence intervals (CIs), estimated with bootstrapping over 1000 resamples. Sample sizes and event counts for each disease example are provided in Supplementary Table 3.

Given that even modest improvements in the C-index at the population level can substantially affect individual risk predictions, we also evaluated the model performance using continuous net reclassification improvement (NRI), which has been shown to provide complementary information on risk model performance [21, 22]. The continuous NRI quantifies the extent to which the model appropriately increases the predicted probabilities for subjects who experience events and decreases them for those who do not. This metric is particularly useful in the absence of established clinical thresholds for high-risk groups, as it quantifies the improvement in risk prediction without relying on predefined risk cutoffs and thus facilitates comparisons across different diseases.

In terms of the continuous NRI, *survivalFM* yielded significantly improved resclassification in 39 out of 40 (97.5%) of the studied examples, with a mean continuous NRI of 37%. Therefore, despite the relatively modest improvement magnitudes in the C-indices, the continuous NRI indicated notable positive changes in individual risk predictions. For instance, type 2 diabetes modelled using clinical biochemistry and blood counts data demonstrated the highest continuous net reclassification improvement of 94% (95% CI 92%-96%), corresponding to 33% (95% CI 32%-35%) of events and 61% (95% CI 60%-61%) of non-events having improved risk estimates (Supplementary Figure 6). Similarly, chronic liver disease models demonstrated notable improvements across all data modalities, with continuous net reclassification improvements ranging between 17%-88%.

These findings suggest that the interaction terms carry additional predictive information across various disease and data modalities and *survivalFM* can model this residual contribution. While the extent of improvement varied depending on the specific disease and dataset under study, improvements were consistently observed across multiple disease areas and data types.

### Disease-specific interaction profiles

A key advantage of *survivalFM* is that despite introducing a more complex layer of non-linearity through the interaction terms, it still maintains interpretability and transparency of how the model predictions are made. Analysis of the estimated interaction effects revealed that in many cases there was a diverse interaction landscape contributing to these predictions, demonstrating that the observed performance gains are likely to stem from the cumulative benefit of many small interaction effects rather than a few prominent ones (examples shown in Supplementary Figures 7-11). Here, we will highlight a few examples with the most notable performance gains.

Inclusion of interaction terms was particularly advantageous in liver-related conditions, such as when predicting alcoholic liver disease or liver fibrosis and cirrhosis using standard risk factors or metabolomic biomarkers. In both liver disease models derived using standard risk factors, among the most prominent interactions were those among different cholesterol measures, cholesterol-lowering medication, and sex (Supplementary Figures 7-8). These results suggest that the joint effects of these risk factors further explain the risk of chronic liver disease outcomes beyond their additive linear effects. The model for alcoholic liver disease also highlighted interactions with white ethnic background, suggesting variation in risk factor profile by ethnicity. Additionally, smoking status was highly weighted both individually and in the interactions, aligning with the earlier research suggesting that smoking may exacerbate the influence of the other risk factors in the development chronic liver diseases [23].

In the case of liver disease models derived using metabolomics biomarkers, both alcoholic liver disease and liver fibrosis and cirrhosis models weighted highly interactions across various amino acids along with their individual effects (Supplementary Figures 9-10). These observations align with previously reported changes in amino acid metabolism related to chronic liver diseases [24, 25], with these results suggesting that the associations of amino acids with the chronic liver disease outcomes are also characterised by complex joint effects. Furthermore, both models emphasized a strong interaction between acetate and glutamine, with acetate having a notably pronounced interaction profile in the model for alcoholic liver disease. Given the known roles of acetate and glutamine in alcohol metabolism and lipid accumulation in the liver [26, 27], these findings indicate that the joint presence of high levels of both these metabolites indicates an even higher risk of chronic liver diseases.

A contrasting example was type 2 diabetes modelled using clinical biochemistry and blood counts data, which obtained the highest observed continuous NRI. Unlike the other examples, analysis of the model coefficients revealed that the model weights were predominantly concentrated around glycated hemoglobin (HbA1c) and its interactions across the other variables (Supplementary Figure 11). The highest interaction weight was attributed to the interaction between HbA1c and glucose, which was negatively weighted despite their positive individual effects. This likely reflects the fact that the simultaneous elevation of both HbA1c and glucose does not increase risk additively but rather relates to them being correlated measures of blood glucose regulation and overall glycemic control. Additionally, the model highlighted positively weighted interactions of HbA1c with age, white ethnicity, and urate levels, indicating these factors together might amplify the risk. In contrast, interactions between HbA1c and reticulocyte count and body mass index were negatively weighted.

### *survivalFM* benefits from large training data sizes

To understand the impact of training data size on model performance and the ability of *survivalFM* to leverage interaction terms, we conducted analyses with models trained on varying-sized subsets of the training data. Throughout these analyses, the test and validation sets were held fixed, allowing us to analyze how changes only in the number of training individuals influence model performance. Figure 3 shows the discriminatory performance of *survivalFM* as a function of the number of training individuals for the input dataset involving standard risk factors (results for the other predictor sets are shown in Supplementary Figures 12-14). These results demonstrate a clear dependency on large sample sizes to uncover predictive interaction terms, with *survivalFM* generally requiring at least 50,000 individuals in training to outperform standard Cox regression. The discriminatory performance of *survivalFM* shows a positive trend and increasing gap to standard Cox regression with increasing sample sizes, although the gains often begin to plateau at the upper end of the sample size range.

**Figure 3:**
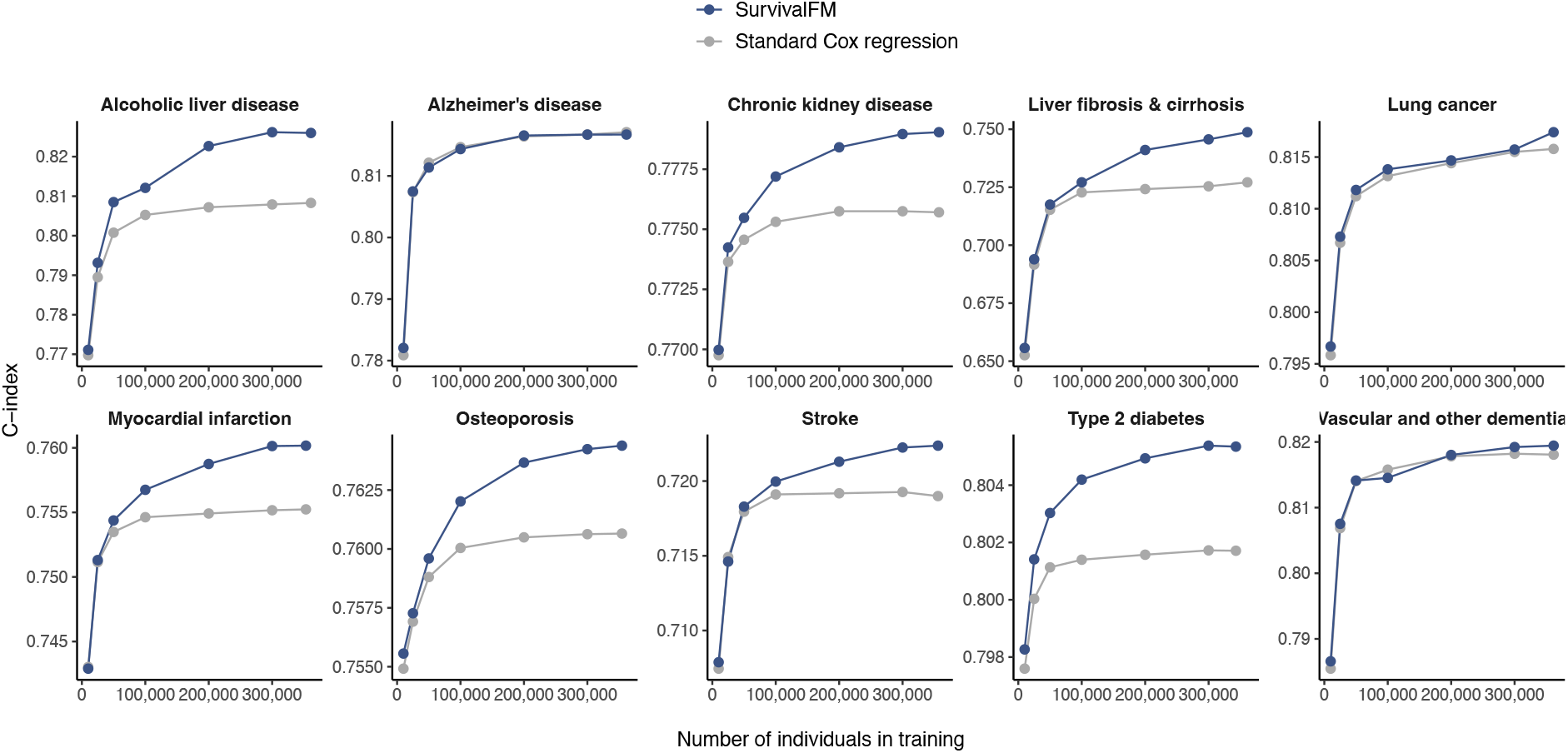
Comprehensive interaction modelling using *survivalFM* benefits from large training data sizes. Impact of the size of the training dataset (x-axis) on the discrimination performance as measured by concordance index (C-index; y-axis), comparing *survivalFM* (blue) to standard Cox regression (gray). Results are shown for the input dataset consisting of standard risk factors. Sample sizes and event counts for each example are provided in Supplementary Table 3.

### *survivalFM* improves prediction performance in a clinical cardiovascular risk prediction scenario

To explore whether comprehensive interaction modeling via *survivalFM* could also refine well-established clinical risk prediction models, we conducted analyses in a clinical CVD risk prediction setting using predictors from the QRISK3 model [5]. QRISK models are Cox proportional hazard models used for predicting the patient’s 10-year risk of CVD, recommended by the healthcare guidelines in the UK. The latest version, QRISK3 from 2017 [5], incorporates a variety of risk factors and comorbidities, along with a set of their interaction terms with age.

We aimed to determine if comprehensive modelling of the interaction terms among the QRISK3 risk factors using *survivalFM* could improve the model’s ability to predict cardiovascular risk. The endpoint was defined as 10-year incidence of composite CVD, including coronary heart disease, ischemic stroke, and transient ischemic attack, and including both fatal and non-fatal events (Supplementary Table 5, Methods). Following the exclusion criteria from the QRISK3 derivation study, we excluded participants with prior CVD diagnoses and those on a cholesterol-lowering medication at the study entry. The baseline characteristics of the study population in this clinical prediction scenario are detailed in Supplementary Table 6.

To ensure a fair comparison of the models, we retrained the QRISK3 model in the UK Biobank considering the same set of risk factors (Methods). As prior research has shown QRISK3 to systematically overestimate CVD risk in the UK Biobank [28], retraining the model ensures an accurate calibration for this cohort. We evaluated three models of increasing complexity: 1) a standard Cox regression model with linear terms only, 2) a Cox regression model incorporating linear terms and age interaction terms from the QRISK3 model, and 3) a *survivalFM* model including linear terms and all potential factorized pairwise interaction terms.

In terms of discrimination performance measured by C-index, *survivalFM* showed statistically significant improvements over the compared models (Figure 4a, Supplementary Table 7). Specifically, it improved the discrimination performance by 0.0018 (95% CI 0.0013-0.0023) over the standard Cox model with linear terms only, and by 0.0014 (95% CI 0.0010-0.0019) over the Cox model including the also the current age interaction terms from QRISK3. Notably, the inclusion of age interaction terms from QRISK3 improved the discrimination performance by 0.0004 (95% CI 0.0000-0.0008) over the model with linear terms only. Hence, modelling the comprehensive interactions using *survivalFM* more than four times improved the discrimination performance gains compared to only incorporating the currently included age interactions.

**Figure 4:**
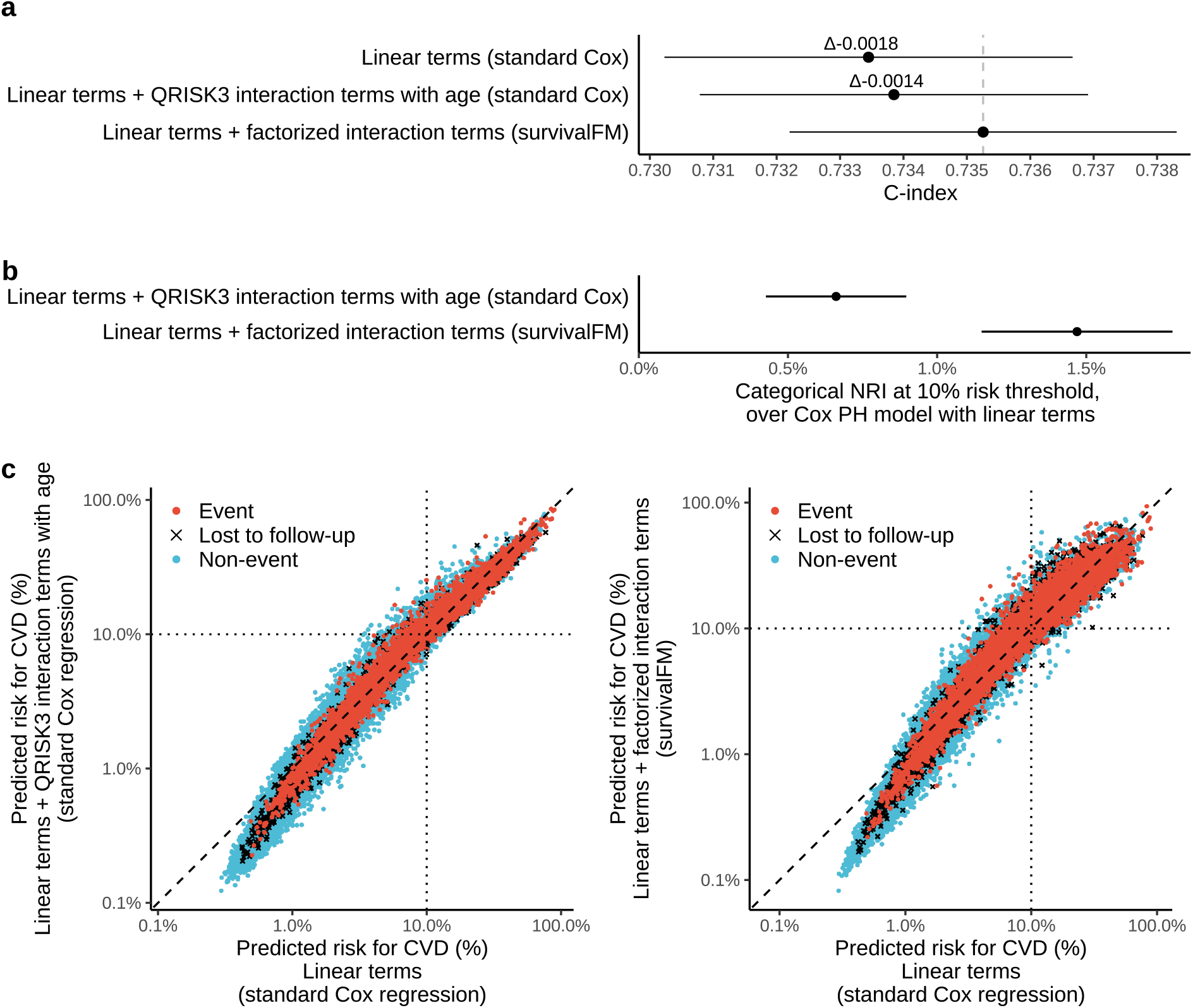
Evaluation of *survivalFM* in a practical clinical cardiovascular risk prediction scenario involving predictors from QRISK3. Performance of the models trained considering QRISK3 predictors for composite cardiovascular disease prediction (N = 344 292 with complete data, 21 534 events). a) Discrimination performance evaluated using concordance index (C-index) for three models: standard Cox regression with linear terms, standard Cox regression with linear terms + age interaction terms from QRISK3 and *survivalFM* model with linear terms + all factorized pairwise interactions. b) Categorical net reclassification improvements (NRI) at 10% absolute risk threshold, as compared to standard Cox model with linear terms. Horizontal error bars denote 95% confidence intervals (CIs), estimated with bootstrapping over 1000 resamples. c) Reclassification plots showing how the inclusion of interaction terms in the more advanced models (y-axis, logarithmic scale) changes individual risk predictions, as compared to a standard linear Cox model (x-axis, logarithmic scale). Black dotted vertical and horizontal lines show the 10% absolute risk threshold for the high risk category.

To further assess how well the models reclassified individuals into appropriate risk categories, we computed categorical net reclassification improvements (NRI) at the guideline recommended 10% absolute risk threshold [29]. Incorporating the currently included age interaction terms from QRISK3 resulted in an overall NRI of 0.66% (95% CI 0.40%-0.93%) compared to the model with linear terms only (Figure 4b). The results for *survivalFM* showed a greater overall NRI of 1.47% (95% CI 1.12%-1.82%), again demonstrating further gains beyond the currently included age interaction terms. *survivalFM* accurately reclassified 3.18% of individuals who experienced an event into the high-risk category, while it inappropriately reclassified a smaller portion of 1.71% of non-events as high-risk (Supplementary Table 7). These improvements are also visible in the reclassification plots (Figure 4c) showing how the individual predictions change with the inclusion of new model terms. All models were well calibrated (Supplementary Figure 15a) and exhibited broadly similar distributions across the risk spectrum (Supplementary Figure 15b).

Analysis of the model coefficients from *survivalFM* revealed a broad array of interactions contributing to the CVD predictions. The ratio of total cholesterol to HDL cholesterol demonstrated the most pronounced interaction profile among all predictor variables (Figure 5). This suggests that the effect of the cholesterol ratio on CVD risk is influenced by the presence of other risk factors. For example, the interaction weight for the cholesterol ratio with prevalent atrial fibrillation was negative, despite both factors having positive individual weights. This suggests that these variables capture partly overlapping aspects of cardiovascular risk. Atrial fibrillation is often associated with a broader cardiovascular risk [30], which could already be reflected in the elevated cholesterol ratio. This may thus imply that when both risk factors are present, they do not independently add to the risk. Comparing the estimated effects for the model terms overlapping between *survivalFM* and the standard Cox regression model with linear and age interaction terms from QRISK3, the shared terms exhibited very similar weights, with correlation of 0.97 between the estimated effects by the two methods (Supplementary Figure 16). This shows that despite the introduction of complex interactions, the fundamental risk associations remain broadly consistent.

**Figure 5:**
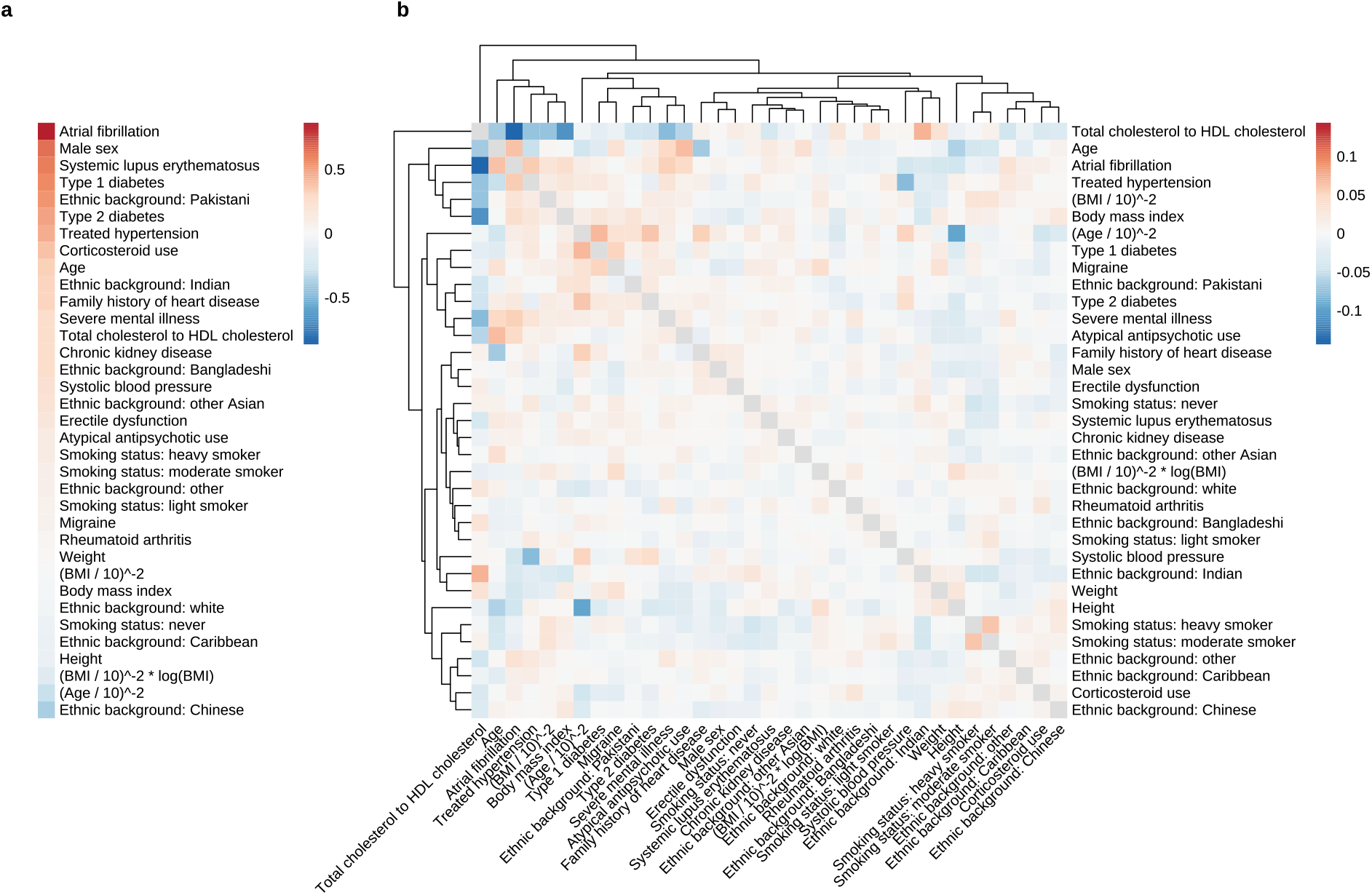
Estimated model coefficients from *survivalFM* model, trained considering the risk factors from QRISK3 for cardiovascular disease risk prediction. The coefficients are shown as the average of the estimated coefficients across the ten models trained during the cross-validation. Estimated coefficients for a) the linear effects ***β*** and b) the interaction effects given by the inner product of the factor vectors *β*_*i,j*_ = ⟨**p**_*i*_, **p**_*j*_⟩. The dendrogram shows a hierarchical clustering of the interaction profiles, using Euclidean distance as the measure of similarity.

## Discussion

Accurate prediction of disease onset and prognosis is essential to realize preventative medicine. In this study, we have introduced *survivalFM*, a new machine learning method for multivariable time-to-event prediction. The method extends the widely used Cox proportional hazards regression by estimating all potential pairwise interaction effects among predictor variables on time-to-event outcomes, such as disease onset. We have shown that estimating these comprehensive interaction effects improves risk prediction and refines individual risk predictions across a range of common diseases, providing more nuanced insights into the interplay among factors underlying disease risk. Since *survivalFM* generalizes to other use cases, we expect this method to find applications in precision medicine and benefit survival modelling in large studies involving many predictors.

Our results from UK Biobank revealed that *survivalFM* can identify predictive interaction terms, which are missed when using standard Cox proportional hazards regression. This ability to uncover predictive interaction terms extended across various disease outcomes and data modalities. Importantly, *survivalFM* consistently matched or surpassed the performance of the standard Cox regression model. This robustness is by design, as *survivalFM* separates linear effects from interaction effects and, by appropriate tuning of model hyperparametrs, can assign negligible weight to non-contributory interaction effects while emphasizing predictive ones. These findings highlight the utility of *survivalFM* in refining risk prediction models across various prediction scenarios, including models derived from traditional clinical predictors and modern omics data types.

Our results further showed that *survivalFM* can add predictive value in practical clinical risk prediction scenarios, such as in CVD risk prediction using predictors from the established QRISK3 model. CVD remains as the leading cause of mortality worldwide [31], making accurate risk stratification critical for healthcare providers to allocate preventive measures effectively. Applying *survivalFM* to QRISK3 risk factors improved both discrimination and reclassification at the clinically recommended 10% risk threshold, more than doubling the performance gains obtained from the current model’s age-related interaction terms alone. For context, while a recent study [18] reported a 1.3% net reclassification improvement by adding a polygenic risk score to a CVD prediction model in a similar scenario involving QRISK3 risk factors, *survivalFM* achieved a comparable improvement by optimizing the use existing clinical variables.

A key strength of *survivalFM* is that despite introducing non-linearity through the comprehensive interaction terms, it maintains interpretability by providing the estimated effects for both the individual terms and the approximated interactions. This is unlike many other advanced machine learning techniques, which often lack transparency. Another advantage of *survivalFM* is a straightforward training process, which only involves optimizing the regularization parameters and setting the rank for factorizing the interaction parameters. We anticipate the accompanying R package will facilitate rapid adoption of the method in other prediction studies.

Interpretation of the trained models suggested that in many cases numerous small interaction effects collectively enhanced the prediction accuracy, highlighting the importance of modeling the entire interaction landscape. However, we also showed that capturing these interaction effects generally requires a large sample size. This can limit the method’s applicability in smaller cohorts and settings with lower sample sizes. Therefore, future studies in adequately powered cohorts are needed to assess the consistency of the identified interactions and gains in prediction accuracy across diverse populations. Whilst large sample size is needed, many biobank initiatives are emerging with clinical and omics data at scale. Our results indicate such initiatives could be used as a base for discovering and replicating comprehensive risk factor interactions that are missed by conventional statistical methods. The generalizable nature of *survivalFM* makes it applicable also to other data modalities than those highlighted in this paper. For instance, comprehensive modelling of interactions across omics data modalities could provide valuable insights into the molecular interplay behind disease risk. Another use case could be studies of protein interaction patterns in relation to disease onset. Recent studies in UK Biobank have demonstrated the strong promise proteomics data in predicting various diseases [32, 33, 34]. Given that proteomics data in UK Biobank comprises around 3 000 measured proteins, the number of potential interactions is in millions. While the current sample size of 50 000 with proteomics data in UK Biobank is at the lower limit for comprehensive interaction modelling, with a sufficient sample size, *survivalFM* could be used to uncover protein interactions predictive of disease onset and potentially provide further insights for personalized treatment strategies.

In conclusion, *survivalFM* provides an advancement in survival analysis, enhancing disease risk prediction by effectively incorporating comprehensive interaction terms. Our findings provide a foundation for future research and translation of risk prediction models, emphasizing the importance of interaction effects in understanding disease development and refining risk prediction models.

## Methods

### *survivalFM*: Extending the proportional hazards model with factorized interaction terms Survival data

Throughout this paper, we assume right-censored survival data. This means that the outcome consists of two variables: the event of interest (here, disease onset) and the time from the beginning of the study period until either to the occurrence of the event, patient loss to follow-up, or end of the duration of follow-up (i.e. right censoring). The survival dataset 𝒟 consists of tuples 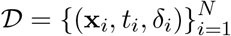, where **x**_*i*_ represents a vector of predictor variables for the individual *i, t*_*i*_ marks the observed time to the event of interest or to the point of censoring, and *δ*_*i*_ is an indicator function which denotes whether *t*_*i*_ corresponds to an actual event occurrence (*δ*_*i*_ = 1) or censored observation (*δ*_*i*_ = 0).

### Model formulation

We base *survivalFM* on the widely used proportional hazards model [1] which relates the time until an event occurs to a set of predictor variables through a hazard function of the from:

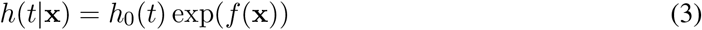

where *h*_0_(*t*) is a shared baseline hazard function that varies over time, and exp(*f* (**x**)) is a partial hazard that describes the effects of the predictor variables on the baseline hazard. In the standard formulation of the Cox proportional hazards model, the partial hazard exp(*f* (**x**)) is assumed to be parametrized by a linear combination of the variables of the individual, *f* (**x**) = ***β***^⊤^**x**, with ***β*** representing the coefficients or parameters of the model assigning weights to the individual variables **x**_*i*_.

In this study, in addition to the individual effects of the variables, we propose to add an approximation of all pairwise interaction terms using a factorized parametrization of the coefficients, following the approach originally introduced along with factorization machines [11] in the context of recommender systems:

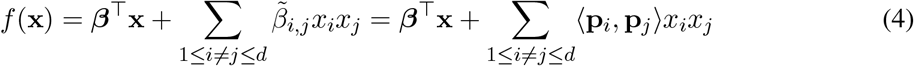

where ⟨·,·⟩ denotes the inner product. The first part contains the linear effects of the predictor variables in the same way as in the standard formulation of the Cox proportional hazards model. The second part contains all pairwise interactions between the predictor variables *x*_*i*_ and *x*_*j*_. However, instead of directly estimating the interaction weights *β*_*i,j*_, the factorized parametrization approximates the coefficients using an inner product between two latent vectors 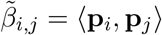. The low-rank factor vectors **p**_*i*_ ∈ ℝ^*k*^ and **p**_*j*_ ∈ ℝ^*k*^ are collected into a parameter matrix **P** ∈ ℝ^*d*×*k*^ (Figure 1a). Rank *k* is a hyperparameter that defines the dimensionality of the factor vectors, and usually the optimal rank of the factorization is much lower than the number of input predictors (*k* ≪ *d*).

### Parameter estimation

Following the standard Cox proportional hazards regression, we estimate the model parameters *θ* using a partial likelihood function *L*(*θ*|𝒟). For each individual who experiences an event at time *t*, their likelihood contribution is the ratio of the hazard of that individual to the cumulative hazard of all other individuals at risk at the same time point, multiplied across all individuals with event occurrence. Formally, this can be expressed as follows:

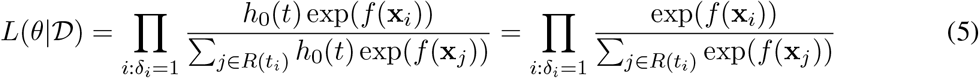

where **x**_*i*_ denotes the vector of predictor variables for an individual *i, t*_*i*_ is the observed event time for individual *i* and *R*(*t*_*i*_) denotes the risk set at time *t*_*i*_. Being in the risk set essentially means that the individual has not had an event yet or that their censoring date has not passed yet. Here, *f* (**x**) corresponds to the log-risk function from eq. (4) containing the individual effects and all pairwise interaction terms in a factorized form. As the baseline hazard function *h*_0_(*t*) is assumed to be shared across all individuals, it cancels out when calculating the partial likelihood, hence eliminating the need for its specification, a key feature of the Cox proportional hazards model rendering it semi-parametric.

To find the optimal parameters *θ* = {***β*, P}**, instead of maximizing the partial likelihood, one can equivalently minimize the negative log-likelihood to obtain a more convenient formulation. Taking the logarithm of the partial likelihood function yields a log-likelihood function of the form:

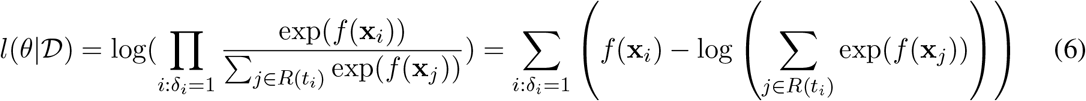

To overcome overfitting in scenarios involving many predictor variables, one can include regularization terms. Here, we consider L2 regularization (Ridge). Hence, the regularized learning problem is given by:

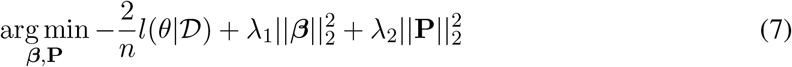

where *λ*_1_ and *λ*_2_ are the regularization parameters for the individual effects and the factorized interactions, respectively. Using separate regularization parameters for the individual effects and the interactions allows for individual penalization of these two parts. The log-likelihood is scaled by a factor of 2*/n* for convenience and to follow the definition from the popular *glmnet* R package [35], used for the standard Cox regression comparison in this study.

Gradient of the negative log-likelihood function *l*(*θ*|𝒟) with respect to the model parameters *θ* = {***β*, P**} is given by:

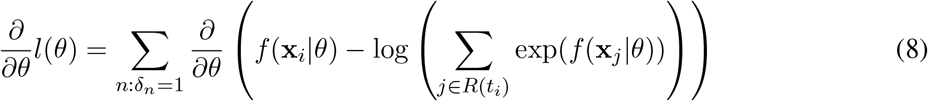

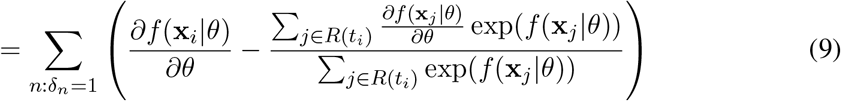

where

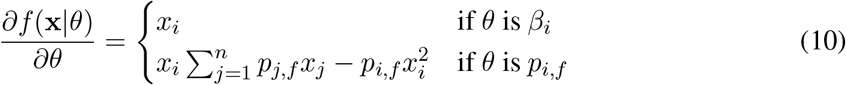

The sum 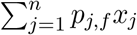 is independent of *i* and thus can be precomputed [11]. In addition, the gradients of the L2 regularization terms are given by 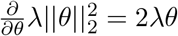.

To solve (7), we use an efficient BFGS (Broyden–Fletcher–Goldfarb–Shanno) quasi-Newton algorithm [36, 37, 38, 39], as implemented in the base R stats package [40]. In contrast to the standard Newton-Raphson method, the BFGS algorithm uses an approximation of the Hessian to determine the search direction. Due to the factorization of the interaction parameters, the number of estimated parameters remains moderate even in the presence of many predictor variables, making the computation of the Hessian approximation feasible. Empirical evidence from our analyses indicated that alternative stochastic gradient descent (SGD)-based optimization methods, commonly employed in machine learning, were not as effective here.

### Study population

The UK Biobank is a comprehensive prospective cohort study serving as a major globally available health research resource. It includes data from approximately half a million participants aged 37-73, representing a sample from the general UK population. The participants were recruited through 22 assessment centers throughout England, Wales, and Scotland between 2006 and 2010. The follow-up is still ongoing. Further details of the study protocol and data collection are available online (https://www.ukbiobank.ac.uk/media/gnkeyh2q/study-rationale.pdf) and in the literature [41]. The UK Biobank study was approved by the North West Multi-Centre Research Ethics Committee and all participants provided written informed consent. In this study, the data was accessed under UK Biobank project ID 147811.

### Predictor variable sets

#### Standard risk factors

As standard risk factors, we included predictors from the ASCVD risk estimator plus [42, 12], which are also commonly featured in other primary prevention tools. These demographic and cardiovascular risk factors have been shown to be predictive of diseases beyond CVD [13, 14, 15]. These included age, sex, ethnic background, systolic and diastolic blood pressure, total, HDL, and LDL cholesterol, smoking status, prevalent type 2 diabetes (excluded from the analyses related to type 2 diabetes), hypertension, and cholesterol-lowering treatment, further detailed in Supplementary Table 8. This data was extracted from the data collected at the study’s initial recruitment visit. Prevalent diabetes status was extracted from primary care records, hospital episode statistics, and self-reported conditions during the initial assessment. These standard risk factors were included in all models trained.

#### Clinical biochemistry and blood counts

A comprehensive set of clinical biochemistry measures were provided by UK Biobank for blood samples taken at the initial recruitment visit and have been previously described in the literature [43, 44]. These included hematologic markers (complete blood counts, white blood cell populations and reticulocytes) and a wide range of blood biochemistry measures covering established risk factors, diagnostic biomarkers and other chracterisation of phenotypes, such as measures for renal and liver function. Nucleated blood cell counts were excluded from our analyses due to over 99% of the cohort having these recorded as missing or zero. We also excluded estradiol, rheumatoid factor and lipoprotein (a), due to a large portion of the cohort (>20%) having these recorded as missing or under the limit of detection. The blood sample handling and storage protocol has been previously described in the literature [45]. A complete list of the included variables is provided in Supplementary Table 9.

#### Metabolomics biomarkers

The metabolomics data included 168 lipids and metabolites from a high-throughput NMR metabolomics assay, available for the baseline blood samples from approximately 275,000 individuals in the UK Biobank. The metabolite data covers a wide range of small molecules, such as amino acids, inflammation markers and ketones, as well as lipids, lipoproteins and fatty acids. Percentage ratios calculated from these 168 original measures were excluded from our analyses. Details of the metabolite data have been previously described [16]. A complete list of the included metabolomics biomarkers is included in Supplementary Table 10.

#### Polygenic risk scores

The polygenic risk score data included 53 polygenic risk scores (PRS) released by the UK Biobank and described in [17]. These included scores for both disease traits and quantitative traits. In our analyses, we included only the standard PRS set obtained entirely from external genome-wide association study (GWAS) data. As provided in the UK Biobank, the score distributions were already centered at zero across all ancestries using a principal component-based ancestry centering step. A complete list of the included variables is provided in Supplementary Table 11.

#### QRISK3 risk factors

We matched the risk factors from the QRISK3 model with the corresponding variables available in the UK Biobank. These variables were gathered during the baseline assessment visit and included cholesterol levels measured from blood samples and prevalent disease diagnoses obtained from linked hospital records, primary care data, and self-reported conditions. In instances where an exact match for a QRISK3 model risk factor was unavailable in the UK Biobank, the closest equivalent field was utilized. A complete list of the predictors and their corresponding UK Biobank fields is provided in Supplementary Table 12.

### Disease endpoint definitions

For the highlighted examples across different data modalities and 10-year incidence of ten different disease outcomes, each of the outcomes was defined by the earliest occurrence in primary care, hospital episode statistics or death records, using the first occurrences data field from UK Biobank (category 1712). For lung cancer, we additionally included data from the cancer registry. The endpoints were defined based on 3-character ICD-10 codes, detailed in Supplementary Table 2. Participants with a previous diagnosis of the disease under study were excluded from the analysis of each endpoint.

The analysis of QRISK3 predictors focused on a 10-year composite CVD outcome, defined according to the original QRISK3 derivation study [5], including coronary heart disease, ischemic stroke, and transient ischemic attack. The ICD-10 codes used are detailed in Supplementary Table 5. We used the earliest recorded date of cardiovascular disease on any of the three data sources (primary care, hospital episode statistics and death records) as the outcome date, using the first occurrences data field from UK Biobank (category 1712). Participants with a prior CVD diagnosis and those on a cholesterol lowering medication at the start of the study were excluded from the analyses, following the exclusion criteria from the original QRISK derivation study [5].

### Data partitions and preprocessing

The model training and testing was performed using a 10-fold cross-validation approach. In each cross-validation cycle, one of the 10 folds at a time was set aside as a test set, aggregating the remaining partitions to form a training set. From this training set, we randomly selected 20% to use as the validation set. Within each of the 10 cross-validation loops, the individual test set remained untouched throughout model development and the validation set was used for model hyperparameter selection. After selecting the optimal hyperparameters, the validation and training sets were combined to train the final model. All 10 obtained models were then evaluated on their respective test sets, and the results from the test sets were aggregated for the final evaluation.

For data preprocessing, log-normally distributed continuous variables (concerning clinical biochemistry markers, blood counts and metabolomics biomarkers) were log1p-transformed (i.e. taking the logarithm of the given value plus one). Outliers exceeding 4 standard deviations from the mean were winsorized. Continuous variables were scaled to zero mean and unit variance and categorical variables were one-hot encoded. The means and standard deviations used for scaling were calculated from the training set and subsequently applied to the validation and test sets. To maximize sample size for the model training, missing values were imputed within the training set using k-nearest neighbors (kNN) imputation (k = 10). To ensure no data leakage between the training, validation and test sets, imputation was exclusively performed within the training set, sparing the validation and test sets from the imputations. Hence, for the validation and test sets, only individuals with complete data available were included.

### Model hyperparameter tuning

In both the standard linear Cox regression and our proposed *survivalFM* method, we employed L2 (ridge) regularization to control model complexity and prevent overfitting. This requires tuning the regularization parameter *λ*. For *survivalFM*, we allowed differing regularization strengths for the linear (*λ*_1_) and the interaction part (*λ*_2_), to separately control the influence of main effects and interaction effects. All regularization parameters were optimized by considering a series of equally spaced values on a logarithmic scale between {1, 1^-4^}. In addition, *survivalFM* requires setting the rank of the factorization (*k*) for the interaction parameters, which was here set to *k* = 10.

### Analysis of model performance

The standard linear Cox regression models used for the comparisons were trained using the *glmnet* [46, 35] R package. Concordance indices (Harrel’s C-index) were computed using the R package *survival* [47] and net reclassification improvements using the R package *nricens* [48].

Confidence intervals for all metrics were calculated with 1000 bootstrapping iterations. Statistical inferences about differences were based on the distributions of bootstrapped performance difference metrics by considering performances statistically significantly different when the 95% confidence intervals did not overlap zero. All analyses were performed using R version 4.3.1 [40].

## Code availability

The method developed in this study has been made available as an R-package and can be installed from: https://github.com/aalto-ics-kepaco/survivalfm.

## Data availability

UK Biobank data are available to researchers upon application at https://www.ukbiobank.ac.uk/enable-your-research/apply-for-access.

## Author contributions

H.J. conceived the idea and designed the method with input from J.R. H.J. processed the data, wrote the code, performed the analyses, prepared the figures and wrote the manuscript. J.R. supervised the work and contributed to writing the manuscript. Both authors read and approved the final manuscript.

## Acknowledgments

This work was supported by the Research Council of Finland grants 339421 (Machine Learning for Systems Pharmacology, MASF, 2021-2025) and 345802 (AI technologies for interaction prediction in biomedicine, AIB, 2022-2024). The authors acknowledge the computational resources provided by the Aalto Science-IT project.

