## Supplementary Figures for "Machine learning for comprehensive interaction modelling improves disease risk prediction in the UK Biobank"

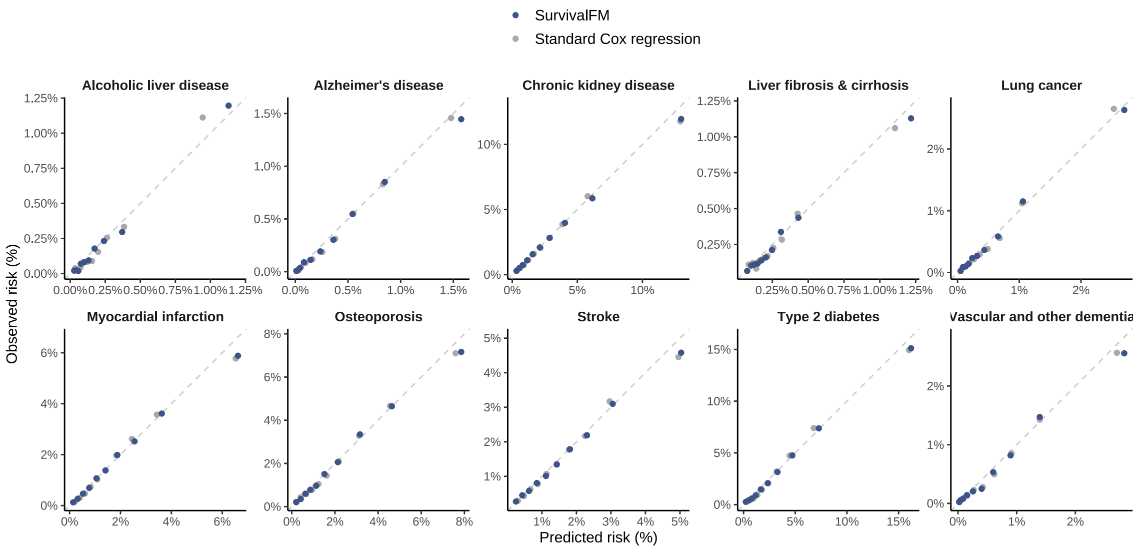


**Supplementary Figure 2:** Calibration of the risk prediction models, trained with the input dataset consisting of standard risk factors. For each model, the observed and predicted event rates are shown for deciles of absolute predicted risk. Sample sizes and event counts are provided in Supplementary Table 3.

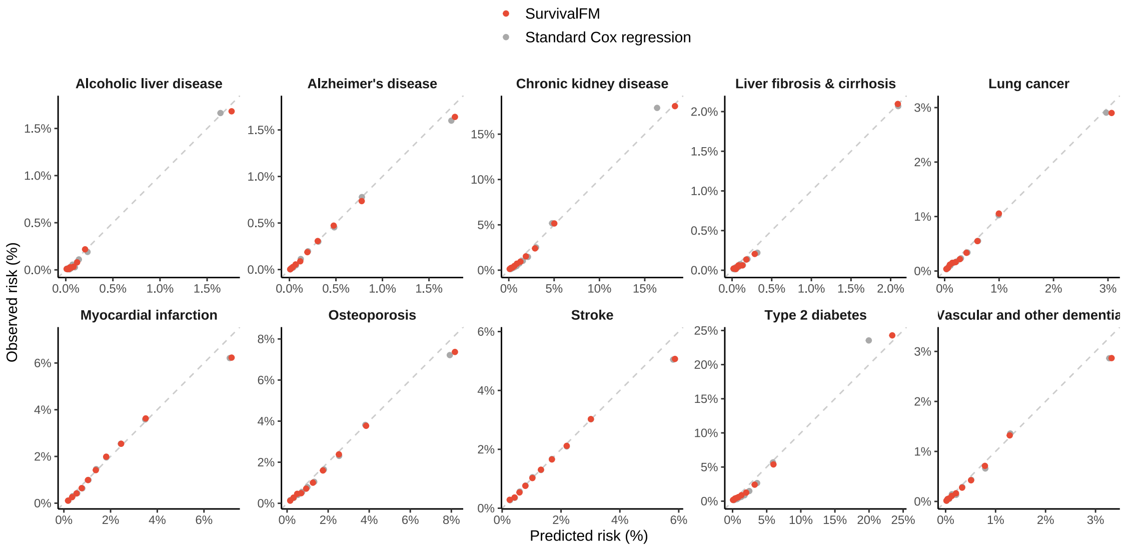


**Supplementary Figure 3:** Calibration of the risk prediction models, trained with the input dataset consisting of clinical biochemistry markers and blood counts. For each model, the observed and predicted event rates are shown for deciles of absolute predicted risk. Sample sizes and event counts are provided in Supplementary Table 3.

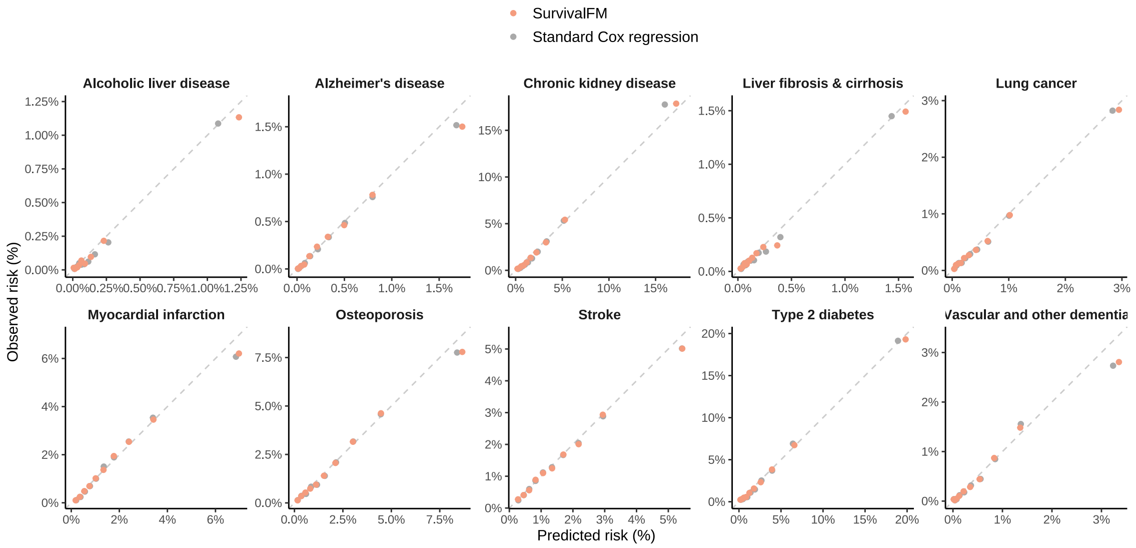


**Supplementary Figure 4:** Calibration of the risk prediction models, trained with the input dataset consisting of metabolic biomarkers. For each model, the observed and predicted event rates are shown for deciles of absolute predicted risk. Sample sizes and event counts are provided in Supplementary Table 3.

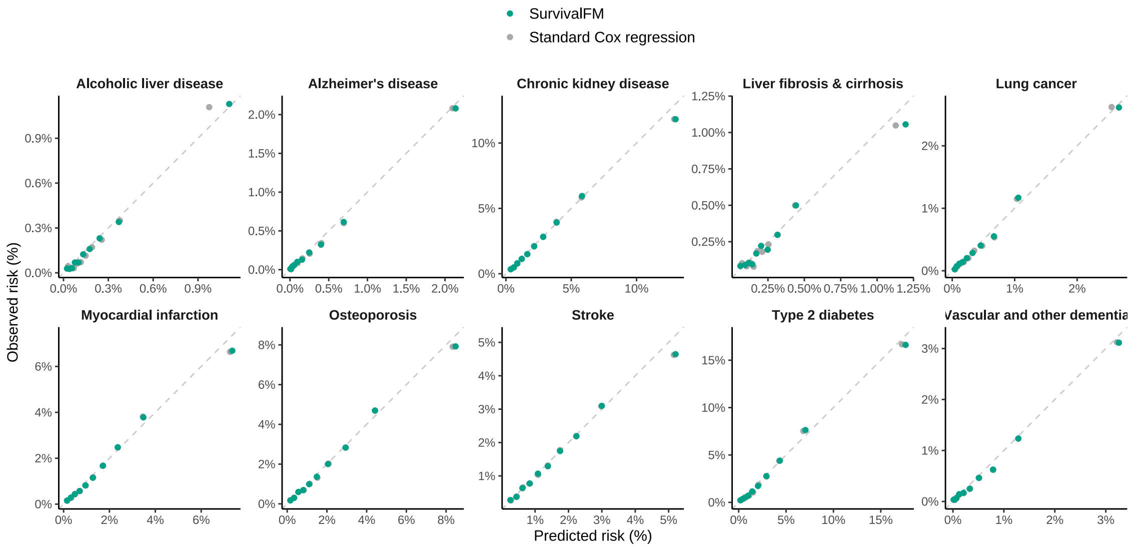


**Supplementary Figure 5:** Calibration of the risk prediction models, trained with the input dataset consisting of polygenic risk scores. For each model, the observed and predicted event rates are shown for deciles of absolute predicted risk. Sample sizes and event counts are provided in Supplementary Table 3.

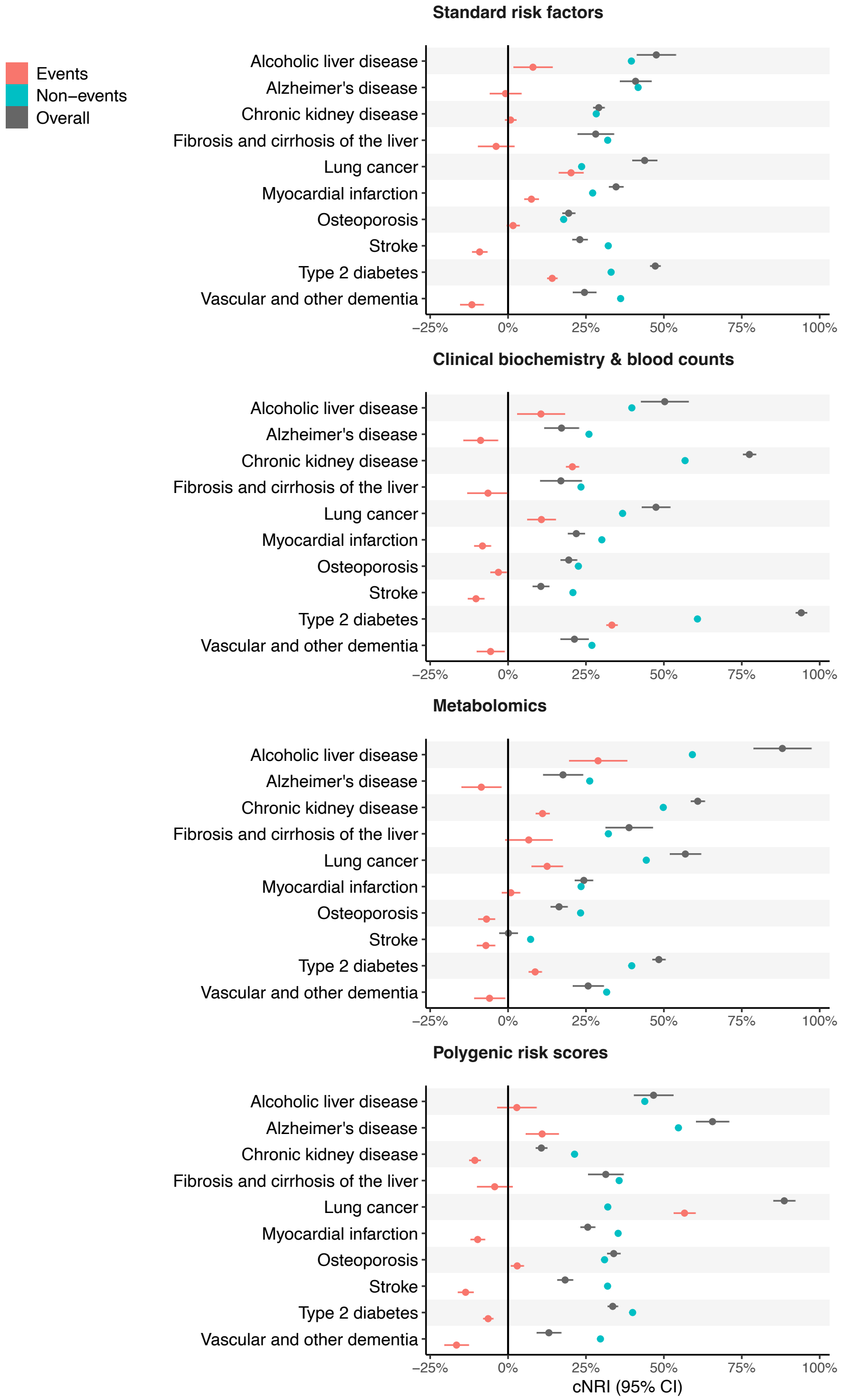


**Supplementary Figure 6:** Continuous net reclassification improvements (cNRI; gray), separated by events (red) and non-events (blue). Results are shown for ten disease examples (y-axis) across four data modalities: standard risk factors (included in all models), clinical biochemistry and blood counts, metabolomics, and polygenic risk scores. Horizontal error bars denote 95% confidence intervals (CIs), estimated with bootsrapping over 1000 resamples. Sample sizes and event counts are provided in Supplementary Table 3.

**Fibrosis and cirrhosis of the liver**

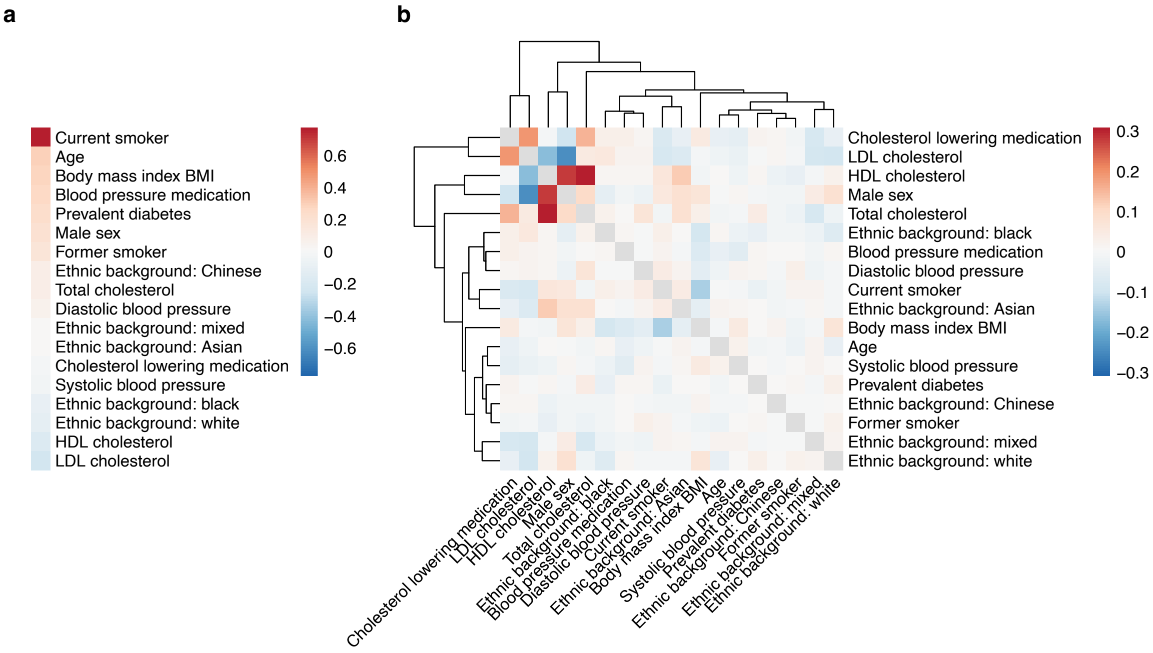


**Supplementary Figure 7:** Coefficients estimated by *survivalFM* for liver fibrosis and cirrhosis, considering the input dataset consisting of standard risk factors. The coefficients are shown as the average of the estimated coefficients across the ten models trained during the cross-validation. Estimated coefficients for a) the linear effects $\boldsymbol{\beta}$ and b) the interaction effects given by the inner product of the factor vectors $\beta_{i,j} = \langle \mathbf{p}_i, \mathbf{p}_j \rangle$. The dendrogram shows a hierarchical clustering of the interaction profiles, using Euclidean distance as the measure of similarity.

**Alcoholic liver disease**

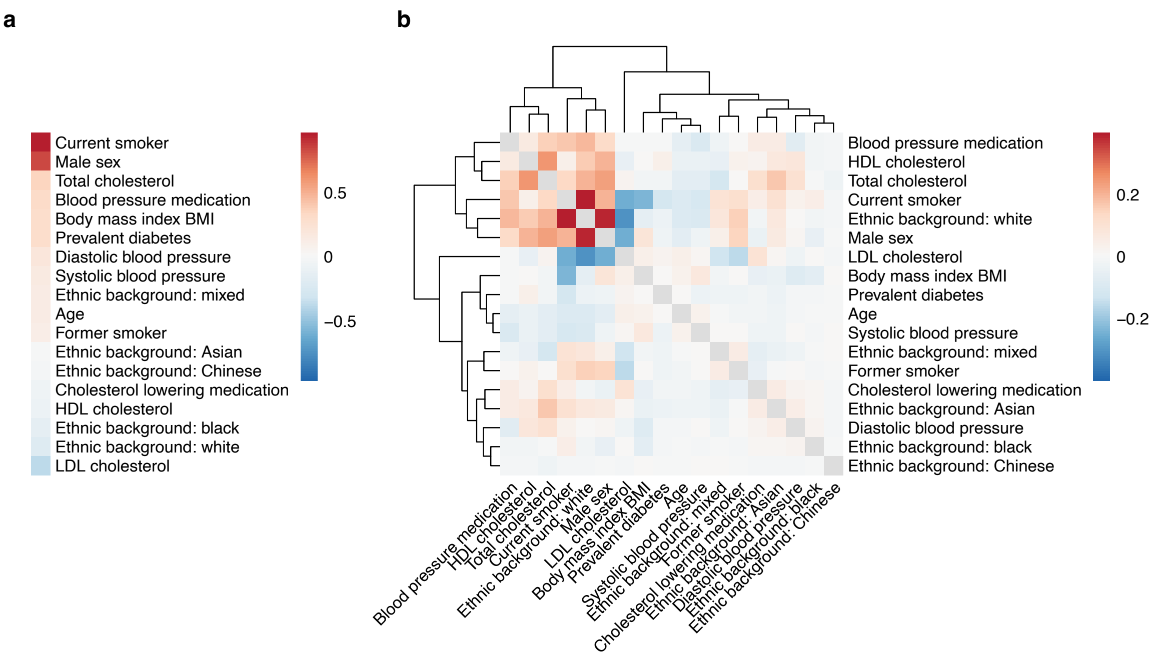


**Supplementary Figure 8:** Coefficients estimated by *survivalFM* for alcoholic liver disease, considering the input dataset consisting of standard risk factors. The coefficients are shown as the average of the estimated coefficients across the ten models trained during the cross-validation. Estimated coefficients for a) the linear effects $\boldsymbol{\beta}$ and b) the interaction effects given by the inner product of the factor vectors $\beta_{i,j} = \langle \mathbf{p}_i, \mathbf{p}_j \rangle$. The dendrogram shows a hierarchical clustering of the interaction profiles, using Euclidean distance as the measure of similarity.

Fibrosis and cirrhosis of the liver

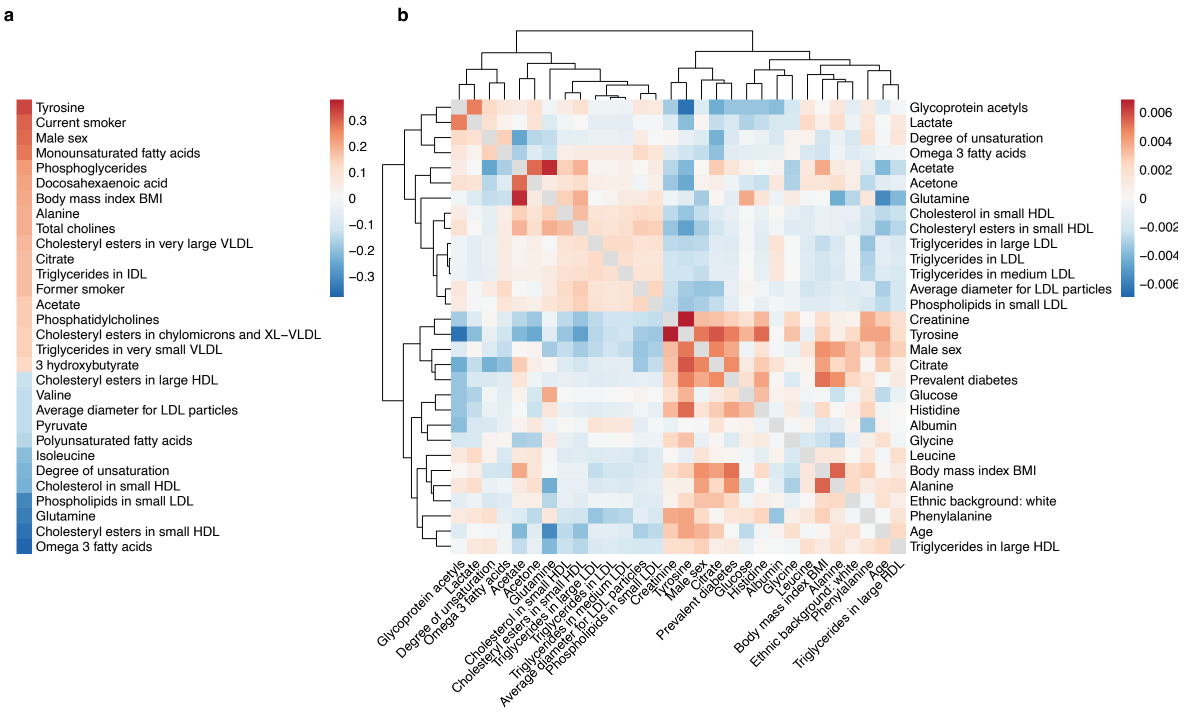


**Supplementary Figure 9:** Coefficients estimated by *survivalFM* for liver fibrosis and cirrhosis, considering the input dataset consisting of metabolic biomarkers. The coefficients are shown as the average of the estimated coefficients across the ten models trained during the cross-validation. Estimated coefficients for a) the linear effects $\boldsymbol{\beta}$ and b) the interaction effects given by the inner product of the factor vectors $\beta_{i,j} = \langle \mathbf{p}_i, \mathbf{p}_j \rangle$. The dendrogram shows a hierarchical clustering of the interaction profiles, using Euclidean distance as the measure of similarity. To facilitate visualization, only top 30 predictors are shown, based on the magnitude of the linear effects (in panel a) and the sum of the magnitudes of the interaction effects (in panel b).

**Alcoholic liver disease**

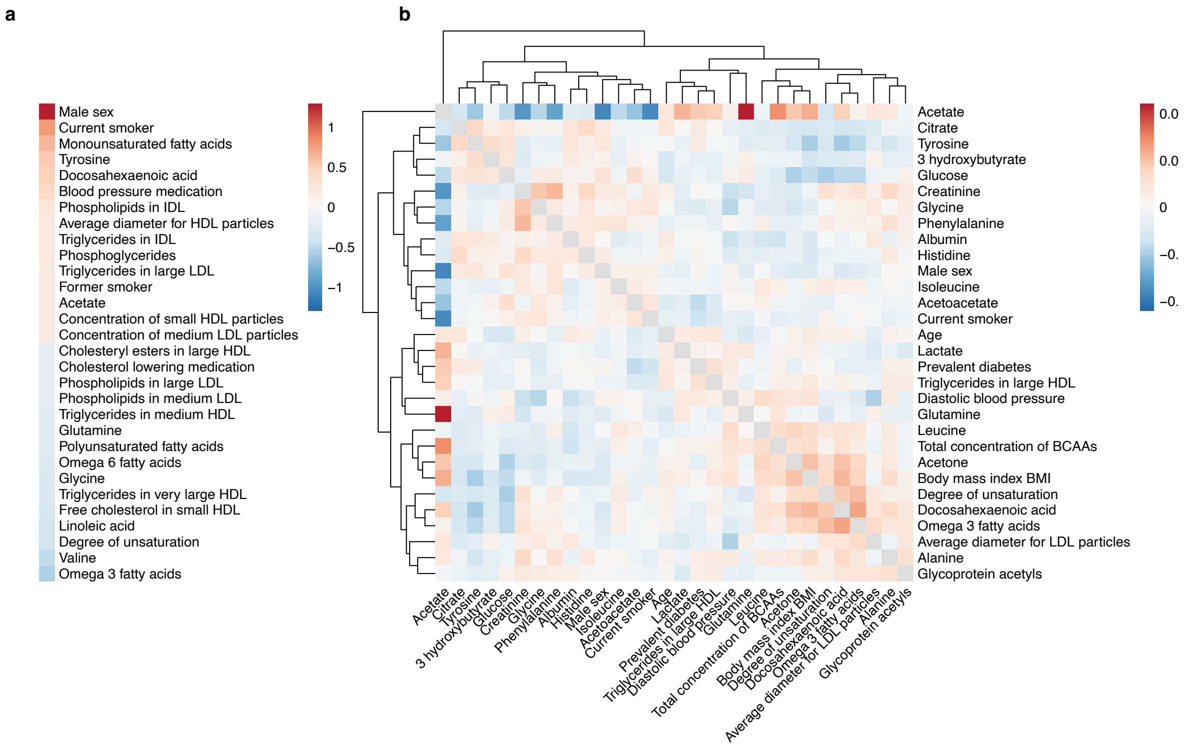


**Supplementary Figure 10:** Coefficients estimated by *survivalFM* for alcoholic liver disease, considering the input dataset consisting of metabolic biomarkers. The coefficients are shown as the average of the estimated coefficients across the ten models trained during the cross-validation. Estimated coefficients for a) the linear effects $\boldsymbol{\beta}$ and b) the interaction effects given by the inner product of the factor vectors $\beta_{i,j} = \langle \mathbf{p}_i, \mathbf{p}_j \rangle$. The dendrogram shows a hierarchical clustering of the interaction profiles, using Euclidean distance as the measure of similarity. To facilitate visualization, only top 30 predictors are shown, based on the magnitude of the linear effects (in panel a) and the sum of the magnitudes of the interaction effects (in panel b).

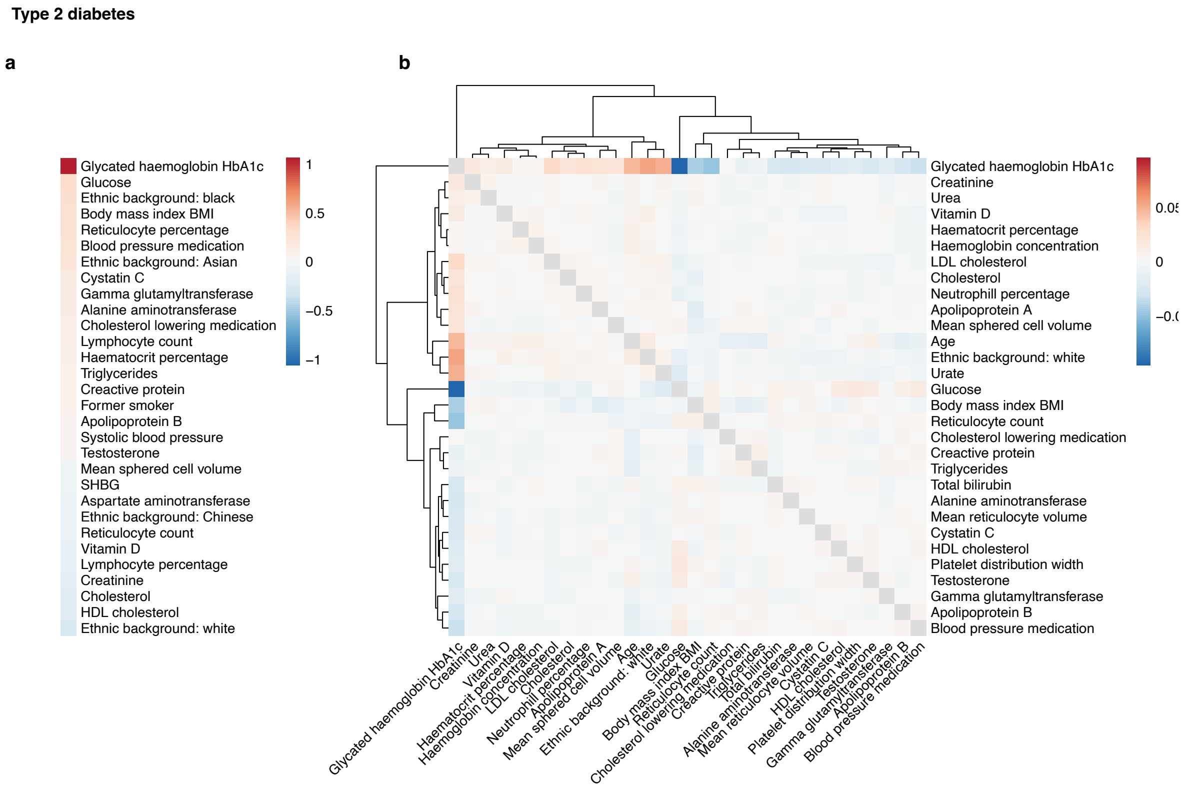


**Supplementary Figure 11:** Coefficients estimated by *survivalFM* for type 2 diabetes, considering the input dataset consisting of clinical biochemistry markers and blood counts. The coefficients are shown as the average of the estimated coefficients across the ten models trained during the cross-validation. Estimated coefficients for a) the linear effects $\boldsymbol{\beta}$ and b) the interaction effects given by the inner product of the factor vectors $\beta_{i,j} = \langle \mathbf{p}_i, \mathbf{p}_j \rangle$. The dendrogram shows a hierarchical clustering of the interaction profiles, using Euclidean distance as the measure of similarity. To facilitate visualization, only top 30 predictors are shown, based on the magnitude of the linear effects (in panel a) and the sum of the magnitudes of the interaction effects (in panel b).

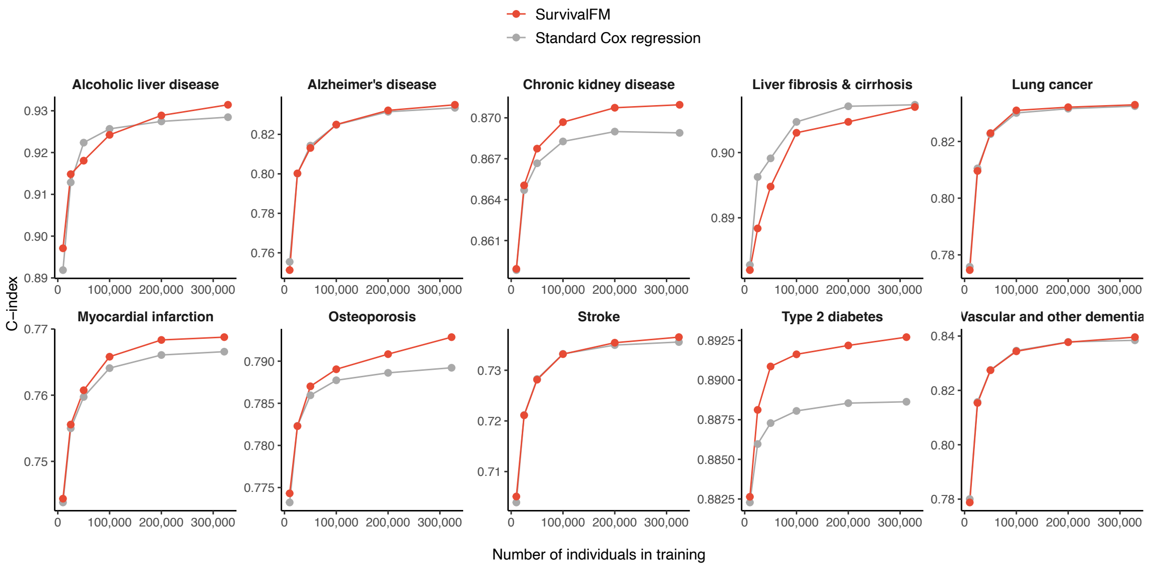


**Supplementary Figure 12:** Impact of the size of the training dataset (x-axis) on the discrimination performance as measured by C-index (y-axis), for the clinical biochemistry & blood counts dataset. *survivalFM* is shown in red, standard Cox regression in gray.

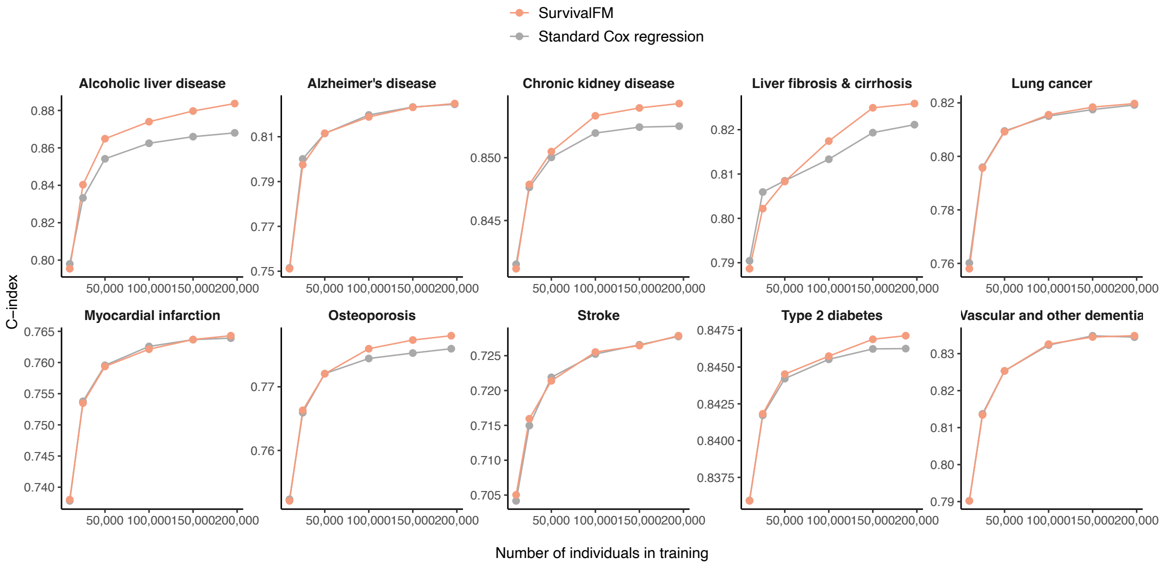


**Supplementary Figure 13:** Impact of the size of the training dataset (x-axis) on the discrimination performance as measured by C-index (y-axis), for the metabolomics dataset. *survivalFM* is shown in orange, standard Cox regression in gray.

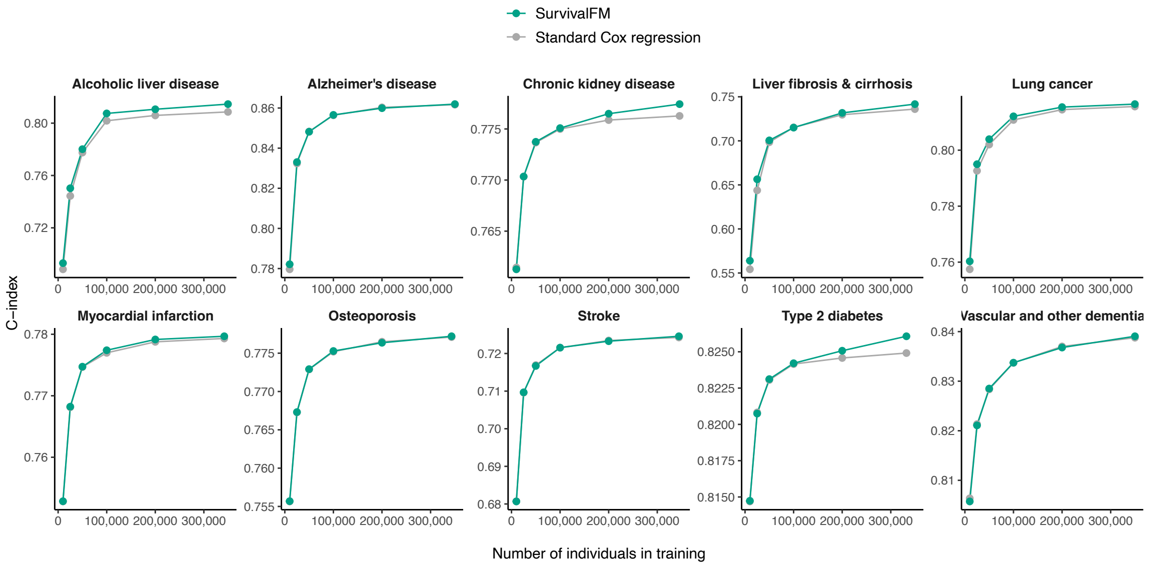


**Supplementary Figure 14:** Impact of the size of the training dataset (x-axis) on the discrimination performance as measured by C-index (y-axis), for the polygenic risk score dataset. *survivalFM* is shown in green, standard Cox regression in gray.

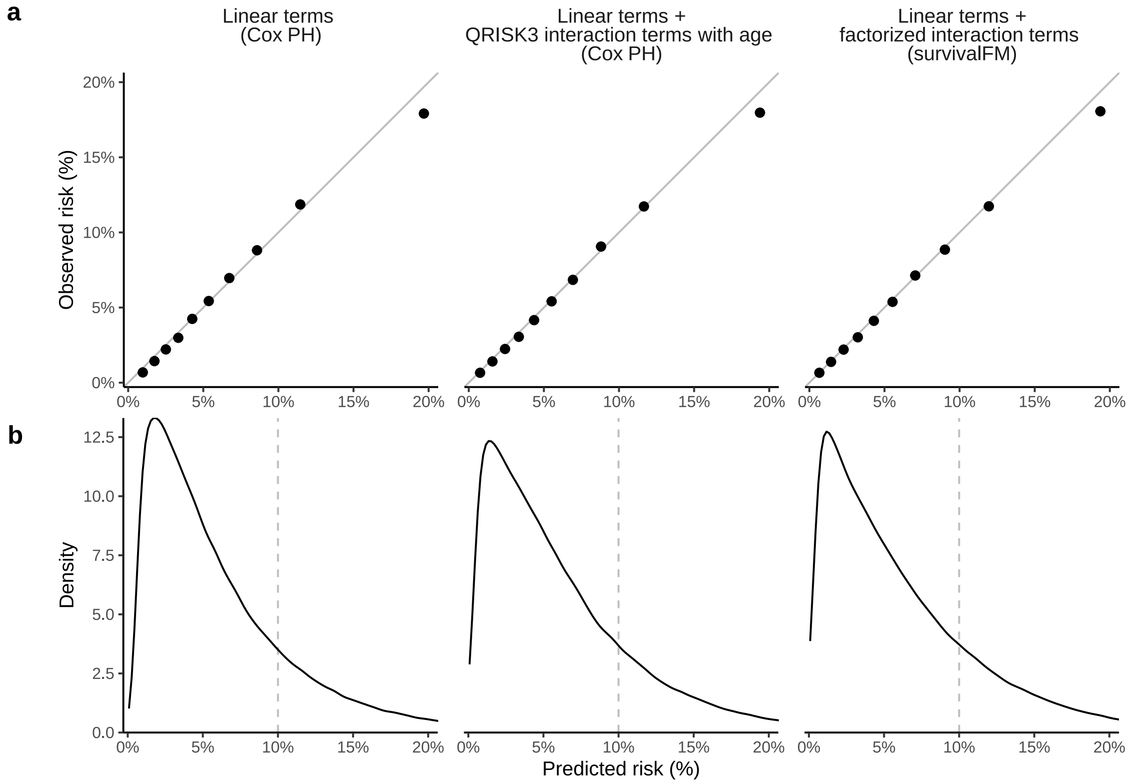


**Supplementary Figure 15:** c) Calibration curves comparing the predicted and observed risks from the analyses involving QRISK3 predictors for the three evaluated models: 1) standard Cox model with linear terms, 2) standard Cox model with linear terms + QRISK3 interaction terms with age and 3) *survivalFM* model with linear terms + all factorized pairwise interaction terms. d) Distributions of predicted risk probabilities.

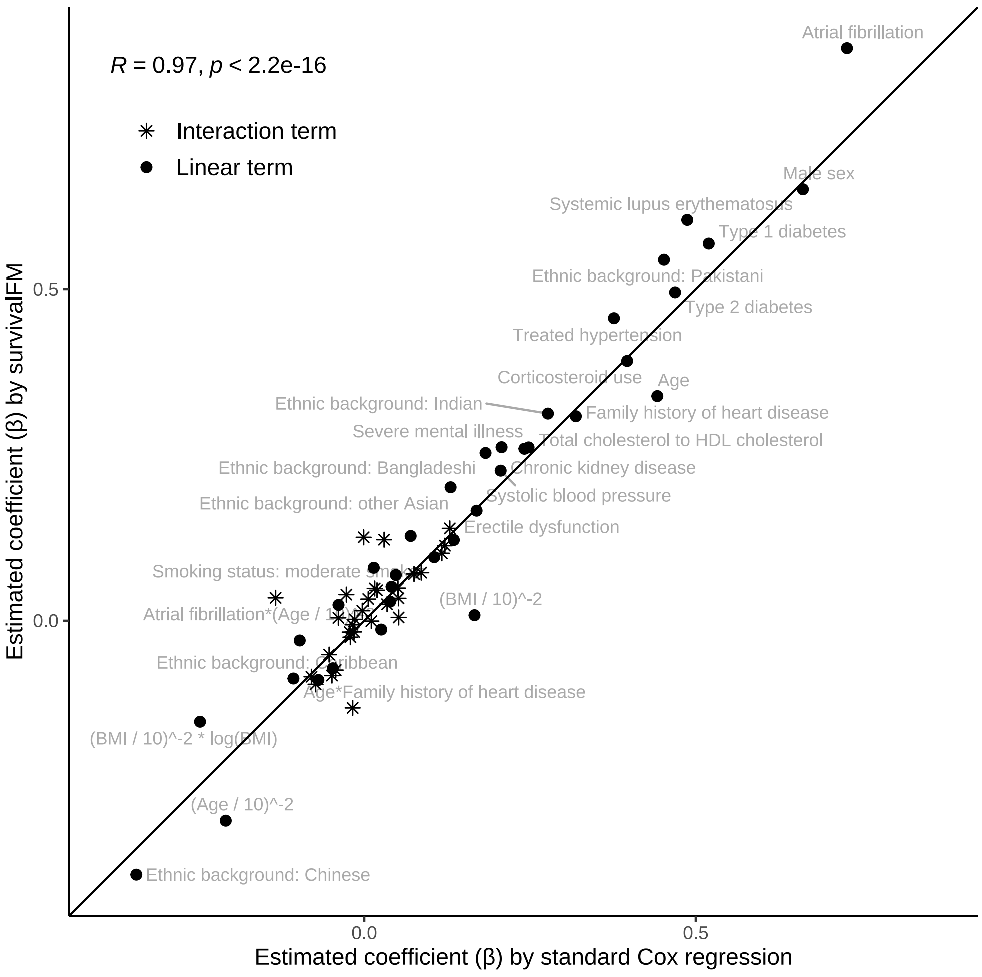


**Supplementary Figure 16:** Model coefficients estimated by standard Cox regression (including linear terms and age interaction terms from QRISK3) vs. the coefficients estimated by *survivalFM* (including linear terms and factorized interaction terms across all input covariate pairs), shown for overlapping terms shared between the two models, in predicting cardiovascular disease using the predictors from QRISK3.
